# The value of fetal placental ratio and placental efficiency in term human pregnancy and complications

**DOI:** 10.1101/2023.02.17.23286091

**Authors:** Peilin Zhang

## Abstract

**Background:** Fetal birth weight and placental weight have been extensively studied and used for clinical assessment of fetal development and maternal health. The ratio of fetal and placental weight as a tool for clinical use in human pregnancy is less studied. We compared the fetal birth weight, placental weight and fetal placental ratio in term pregnancy to see if fetal and placental ratio is useful in assessment of maternal health and pregnancy complication as well as fetal growth and development in singleton pregnancy.

**Material and methods:** We have collected the fetal birth data, maternal pregnancy data and placental pathology data from March 2000 to November 2021 in a single urban hospital. We compared the fetal birth weight, placental weight and fetal placental ratio in assessment of fetal growth, maternal pregnancy complications, and placental pathology with special emphasis on the role of fetal placental ratio.

**Results:** A total 3302 pairs of neonates and placentas from term singleton pregnancy were reviewed and fetal birth weight and placental weight were moderately correlated with Pearman’s correlation coefficiency R=0.66. Fetal placental ratio as a proxy of placental efficiency was significantly associated with various pregnancy complications and placental pathology, and these associations were different from those of fetal birth weight or placental weight alone. High placental efficiency (90 percentile or greater) was associated with ethnic White, SARS-CoV2 status, category 2 fetal heart tracing and maternal inflammatory response in placenta while low placental efficiency (less than 10 percentile) was associated with ethnic Black, Asian and Hispanic, preeclampsia/pregnancy induced hypertension and gestational diabetes mellitus.

**Conclusion:** Fetal placental ratio was shown to be a useful indicator different from fetal birth weight and placental weight alone. Maternal and environmental factors were shown to have differential effects on fetal and placental growth. Understanding the mechanism of differential fetal and placental growth will help better manage the clinical relevant conditions such as IUGR and macrosomia.

## Introduction

Fetal birth weight at term is an important measure for neonatal morbidity and mortality, and it is critical for neonatal growth and development immediately after birth and later in life [1]. Fetal birth weight is influenced by many factors including maternal nutrition, fetal genetics, fetal sex and placental development in utero, and variations of fetal birth weight consequently lead to clinically relevant conditions such as macrosomia or intrauterine growth restriction (IUGR) [2]. Placental growth and development is generally correlated with fetal growth throughout pregnancy as measured by the fetal birth weight and the placental weight at term, but this correlation varies in early and late stages of pregnancy [3]. Placental weight can be influenced by both maternal and fetal circulation, intrauterine location of the placenta, centrality of the umbilical cord insertion on the placental disc as well as maternal or fetal inflammatory response [4, 5]. Both fetal birth weight and placental weight are important information in assessment of fetal health at birth and later in life. Placental efficiency was calculated as fetal placental weight ratio (FPR) representing grams of fetal weight growth per gram of placenta [6, 7]. Placental efficiency reflects nutritional transfer capability of the placenta to support the fetal growth, as well as endocrine functions of the placenta [6, 7]. FPR became interesting as both fetal birth weight and placental weight were associated with chronic diseases in adult life including cardiovascular diseases, hormonal diseases such as diabetes and cancer with emergence of “fetal origin of adult disease” theory [8-12]. Placental efficiency can be demonstrated through various animal models, but in human it is less studied during pregnancy until term or near term after delivery when fetal birth weight and placental weight can be readily measured [13, 14]. As the FPR is apparently affected by either fetal birth weight, or placental weight or both, the value of FPR in clinical setting may be different from that of fetal birth weight or placental weight alone. We sought to examine the FPR in comparison with fetal birth weight and placental weight in regards to pregnancy complications and placental pathology.

## Material and methods

This study followed the Strengthening the Reporting of Observational Studies in Epidemiology (<u>STROBE</u>) reporting guideline and included all term singleton placentas submitted chronologically for pathology examination from March 2020 and November 2021 with the exception of twin or multiple births. Placental examination at our institution is criteria-based and performed according to the standard procedure [15-17]. Placental pathology data, neonatal birth data, maternal racial and ethnic data and marital status were retrieved from medical records from the hospital medical record system (Cerner Corporation) based on standard national criteria. The placental weight was measured after the fetal membrane and umbilical cord were trimmed [15]. Marital status was listed as married, single, divorced, life partner, or others including unknowns and declined to respond. Classification for race and ethnicity included Asian, Hispanic, non-Hispanic Black, and non-Hispanic White categories; responses outside of these categories (ie, “unknown,” “others,” or “declined”) were recorded together as one group. Laboratory tests of white blood cell counts with differentials and blood pressure measurements were from pre-admission tests for delivery only. Only the singleton term pregnancies were included (gestational age of 37 week or over), and preterm pregnancies (less than 37 weeks) were excluded. Statistical analysis was performed by using various programs in R-package including baseline characteristic table and Excel programs. *P*□<□.05 was considered significant. This work was approved by the Institutional Review Board of New York Presbyterian –Brooklyn Methodist Hospital [1592673-1] (approval date 4-13-2020).

## Results

### 1 Placental weight and fetal birth weight were moderately correlated

A total 3302 pairs of term (37 weeks or over) singleton neonates and placentas were included in the study. The mean fetal birth weight was 3300 grams (95% confidence interval 2970 to 3630). The mean trimmed placental weight was 466 grams (95% CI 403.0 to 538.0). The distributions of fetal birth weight and placental weight were shown in Figure 1. In general, the placental weight and fetal birth weight were moderately correlated with the Pearson’s correlation coefficient R to be 0.66 (R=0.66) (Figure 2).

**Figure 1:**
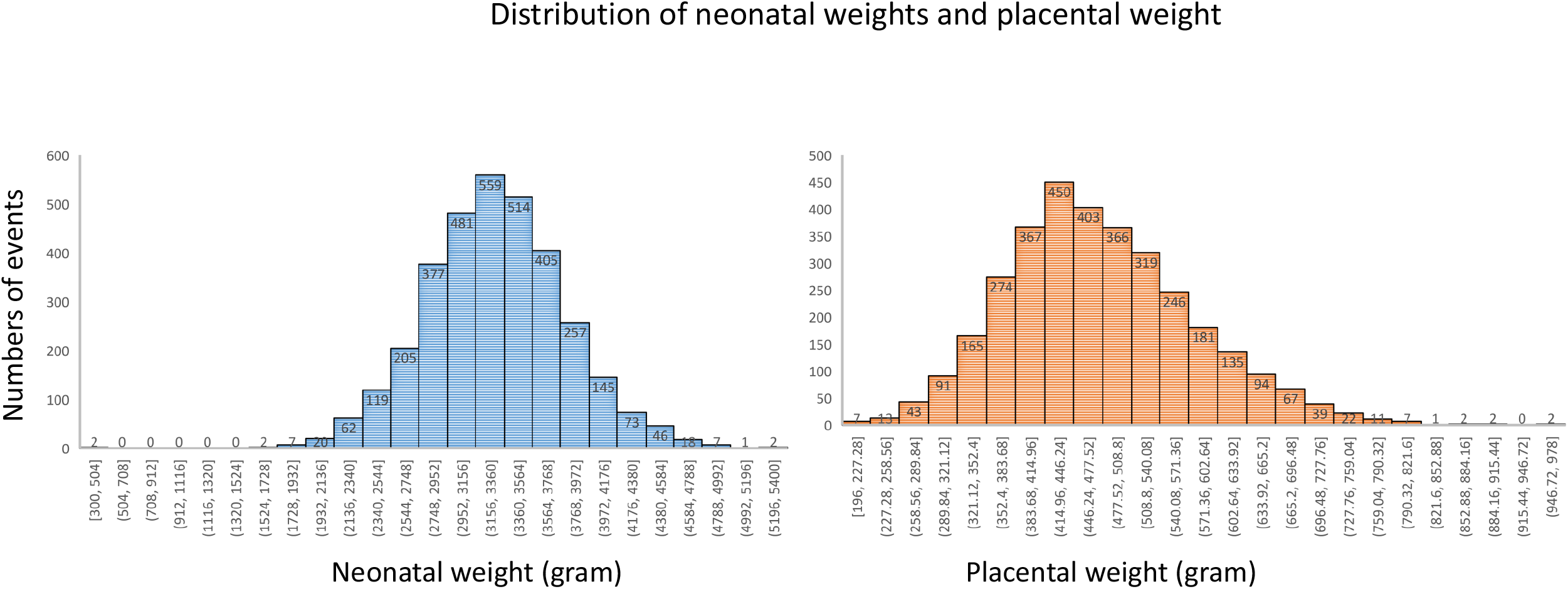
Histograms of fetal birth weight and trimmed placental weight distribution of 3302 term pregnancies. The mean fetal birth weight was 3300 grams (95% CI 2970 to 3630). The mean trimmed placental weight was 466 grams (95% CI 403.0 to 538.0).

**Figure 2:**
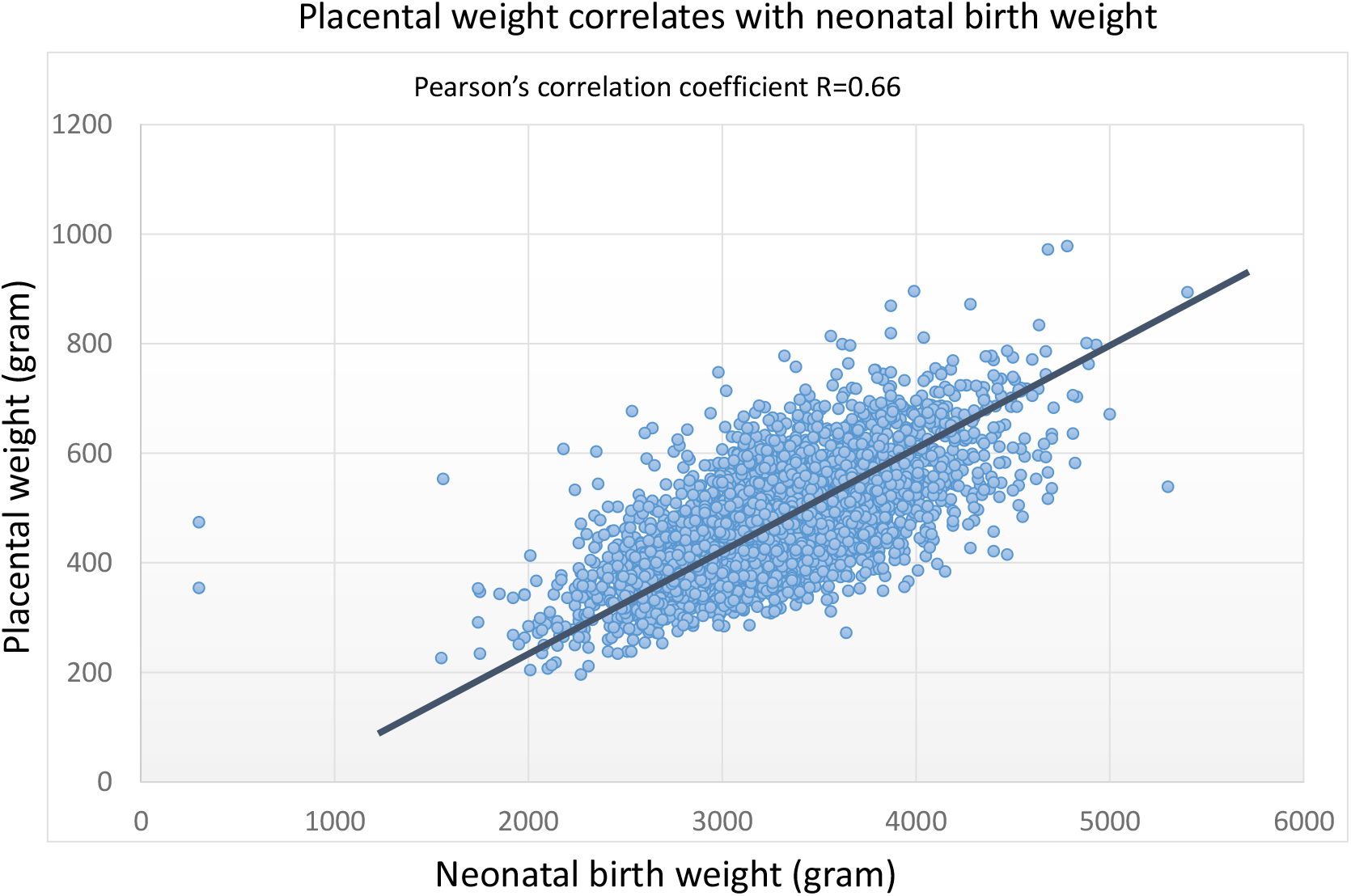
Correlation of fetal birth weight and trimmed placental weight with Pearson’s correlation efficiency R=0.66.

### 2 Fetal placental weight ratio (FPR) and pregnancy complications and placental pathology

The fetal birth weight, placental weight and fetal placental weight ratio (FPR) as well as the percentile ranks were calculated within the Excel table, and the overall FPR in the entire study population was 7.1 (95% CI 6.3 to 7.3) (Table 1). The percentile rank of FPR was divided as three separate groups, the group of 90% or over (mean FPR=9.2, 95% CI 8.9 to 9.7, the top 10%), 10-89% (mean FPR =7.0, 95% CI 6.4 to 7.6), and the group of less than 10% (mean FPR =5.3, 95% CI 5.0 to 5.5, the bottom 10%) (Table 1). The FPR has previously been used as a proxy of placental efficiency (the grams of fetal growth supported by a gram of placenta), and high FPR (90% or over, the top 10%) represents high placental efficiency (HPE), and low FPR (10% or less, the bottom 10%) low placental efficiency (LPE). Although FPR was statistically correlated with fetal birth weight and placental weight, the correlation was non-linear, and the relationships between the fetal birth weight and placental weight were more complexed (Table 1). Notably, ethnic Black showed significantly lower placental efficiency with low FPR in comparison to ethnic White population (Table 1), whereas marital status showed no significant differences in FPR. Maternal obesity was statistically correlated with FPR but the relationship between body mass index (BMI) and FPR was non-linear. Interestingly, the frequency of SARS-CoV2 positive status was found significantly higher in high FPR group (Table 1), but Cesarean section delivery, preeclampsia/PIH, GDM2 were significantly higher in low FPR group. In placental pathology, high FPR group was associated with increased frequency of decidual vasculopathy (both classic type and mixed type), and maternal inflammatory response (MIR) including chronic deciduitis and chronic villitis (Table 1). FPR was not significantly associated with umbilical cord centrality (distance of cord insertion site to placental edge).

**Table 1:**
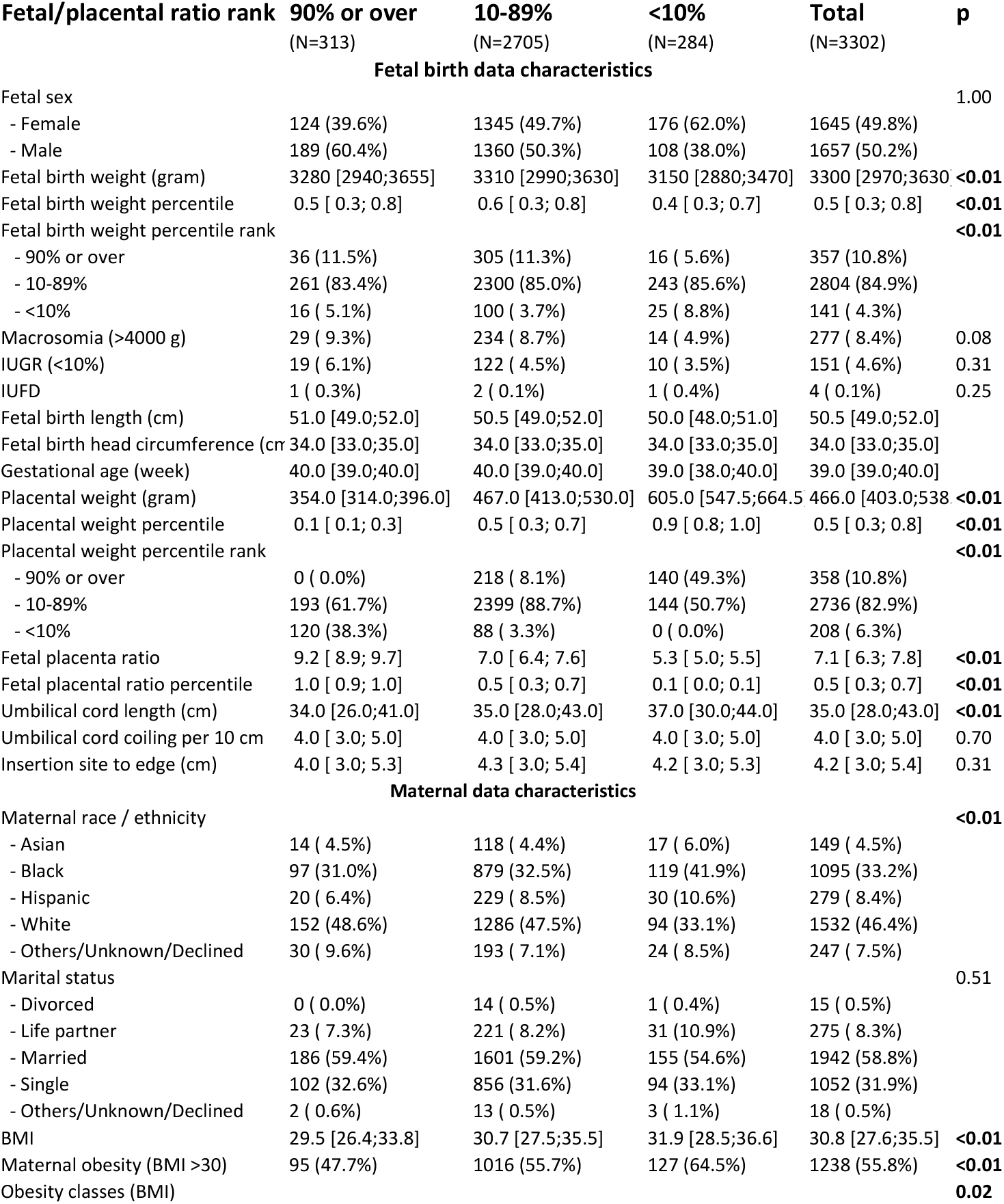
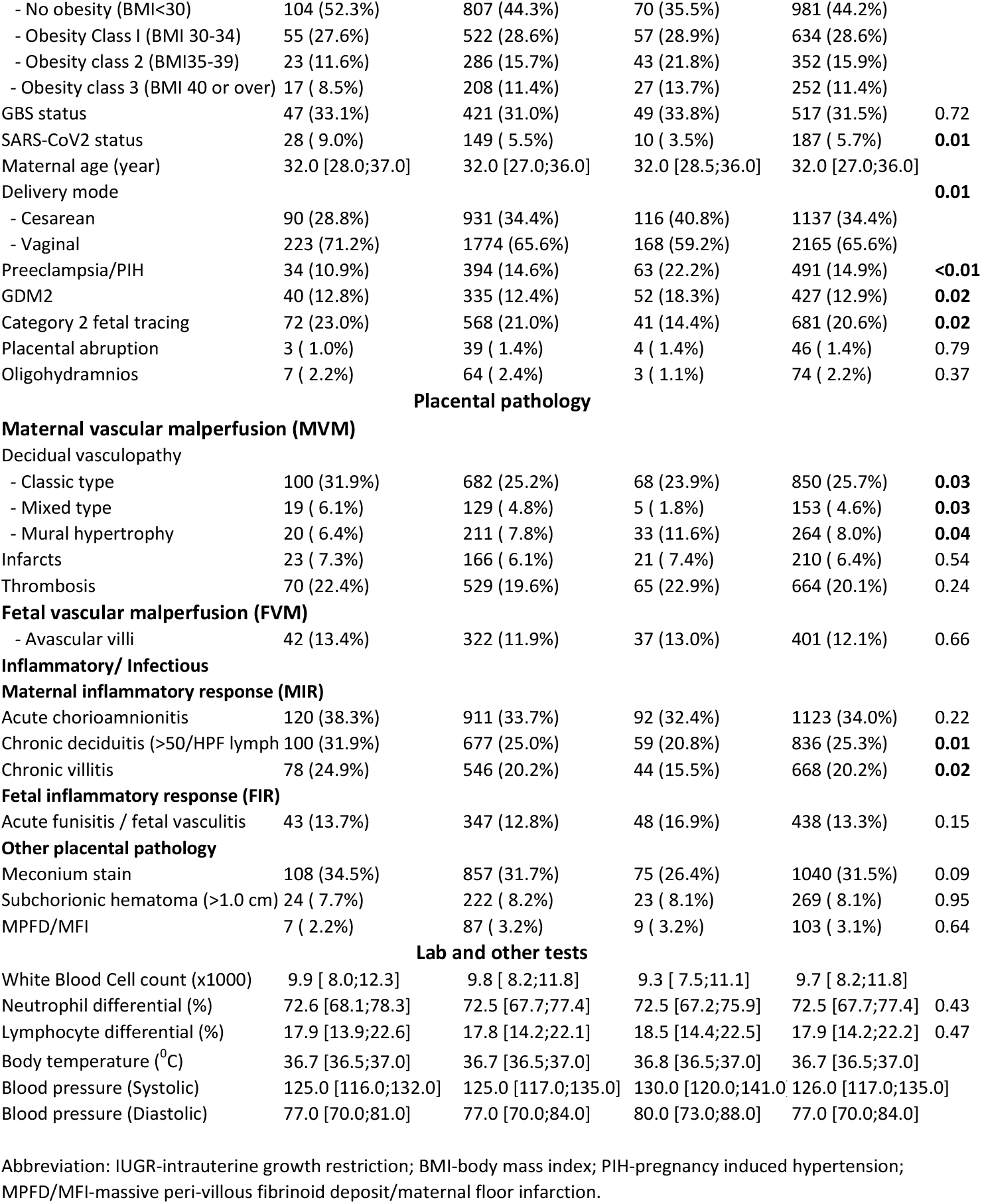
Fetal placental ratio and term pregnancy complications.

| **Fetal/placental ratio rank** | **90% or over**<br>(N=313) | **10-89%**<br>(N=2705) | **<10%**<br>(N=284) | **Total**<br>(N=3302) | **p** |
| --- | --- | --- | --- | --- | --- |
| **Fetal birth data characteristics** |  |  |  |  |  |
| Fetal sex |  |  |  |  | 1.00 |
| - Female | 124 (39.6%) | 1345 (49.7%) | 176 (62.0%) | 1645 (49.8%) |  |
| - Male | 189 (60.4%) | 1360 (50.3%) | 108 (38.0%) | 1657 (50.2%) |  |
| Fetal birth weight (gram) | 3280 [2940;3655] | 3310 [2990;3630] | 3150 [2880;3470] | 3300 [2970;3630] | **<0.01** |
| Fetal birth weight percentile | 0.5 [ 0.3; 0.8] | 0.6 [ 0.3; 0.8] | 0.4 [ 0.3; 0.7] | 0.5 [ 0.3; 0.8] | **<0.01** |
| Fetal birth weight percentile rank |  |  |  |  | **<0.01** |
| - 90% or over | 36 (11.5%) | 305 (11.3%) | 16 ( 5.6%) | 357 (10.8%) |  |
| - 10-89% | 261 (83.4%) | 2300 (85.0%) | 243 (85.6%) | 2804 (84.9%) |  |
| - <10% | 16 ( 5.1%) | 100 ( 3.7%) | 25 ( 8.8%) | 141 ( 4.3%) |  |
| Macrosomia (>4000 g) | 29 ( 9.3%) | 234 ( 8.7%) | 14 ( 4.9%) | 277 ( 8.4%) | 0.08 |
| IUGR (<10%) | 19 ( 6.1%) | 122 ( 4.5%) | 10 ( 3.5%) | 151 ( 4.6%) | 0.31 |
| IUFD | 1 ( 0.3%) | 2 ( 0.1%) | 1 ( 0.4%) | 4 ( 0.1%) | 0.25 |
| Fetal birth length (cm) | 51.0 [49.0;52.0] | 50.5 [49.0;52.0] | 50.0 [48.0;51.0] | 50.5 [49.0;52.0] |  |
| Fetal birth head circumference (cm) | 34.0 [33.0;35.0] | 34.0 [33.0;35.0] | 34.0 [33.0;35.0] | 34.0 [33.0;35.0] |  |
| Gestational age (week) | 40.0 [39.0;40.0] | 40.0 [39.0;40.0] | 39.0 [38.0;40.0] | 39.0 [39.0;40.0] |  |
| Placental weight (gram) | 354.0 [314.0;396.0] | 467.0 [413.0;530.0] | 605.0 [547.5;664.5] | 466.0 [403.0;538.0] | **<0.01** |
| Placental weight percentile | 0.1 [ 0.1; 0.3] | 0.5 [ 0.3; 0.7] | 0.9 [ 0.8; 1.0] | 0.5 [ 0.3; 0.8] | **<0.01** |
| Placental weight percentile rank |  |  |  |  | **<0.01** |
| - 90% or over | 0 ( 0.0%) | 218 ( 8.1%) | 140 (49.3%) | 358 (10.8%) |  |
| - 10-89% | 193 (61.7%) | 2399 (88.7%) | 144 (50.7%) | 2736 (82.9%) |  |
| - <10% | 120 (38.3%) | 88 ( 3.3%) | 0 ( 0.0%) | 208 ( 6.3%) |  |
| Fetal placenta ratio | 9.2 [ 8.9; 9.7] | 7.0 [ 6.4; 7.6] | 5.3 [ 5.0; 5.5] | 7.1 [ 6.3; 7.8] | **<0.01** |
| Fetal placental ratio percentile | 1.0 [ 0.9; 1.0] | 0.5 [ 0.3; 0.7] | 0.1 [ 0.0; 0.1] | 0.5 [ 0.3; 0.7] | **<0.01** |
| Umbilical cord length (cm) | 34.0 [26.0;41.0] | 35.0 [28.0;43.0] | 37.0 [30.0;44.0] | 35.0 [28.0;43.0] | **<0.01** |
| Umbilical cord coiling per 10 cm | 4.0 [ 3.0; 5.0] | 4.0 [ 3.0; 5.0] | 4.0 [ 3.0; 5.0] | 4.0 [ 3.0; 5.0] | 0.70 |
| Insertion site to edge (cm) | 4.0 [ 3.0; 5.3] | 4.3 [ 3.0; 5.4] | 4.2 [ 3.0; 5.3] | 4.2 [ 3.0; 5.4] | 0.31 |
| **Maternal data characteristics** |  |  |  |  |  |
| Maternal race / ethnicity |  |  |  |  | **<0.01** |
| - Asian | 14 ( 4.5%) | 118 ( 4.4%) | 17 ( 6.0%) | 149 ( 4.5%) |  |
| - Black | 97 (31.0%) | 879 (32.5%) | 119 (41.9%) | 1095 (33.2%) |  |
| - Hispanic | 20 ( 6.4%) | 229 ( 8.5%) | 30 (10.6%) | 279 ( 8.4%) |  |
| - White | 152 (48.6%) | 1286 (47.5%) | 94 (33.1%) | 1532 (46.4%) |  |
| - Others/Unknown/Declined | 30 ( 9.6%) | 193 ( 7.1%) | 24 ( 8.5%) | 247 ( 7.5%) |  |
| Marital status |  |  |  |  | 0.51 |
| - Divorced | 0 ( 0.0%) | 14 ( 0.5%) | 1 ( 0.4%) | 15 ( 0.5%) |  |
| - Life partner | 23 ( 7.3%) | 221 ( 8.2%) | 31 (10.9%) | 275 ( 8.3%) |  |
| - Married | 186 (59.4%) | 1601 (59.2%) | 155 (54.6%) | 1942 (58.8%) |  |
| - Single | 102 (32.6%) | 856 (31.6%) | 94 (33.1%) | 1052 (31.9%) |  |
| - Others/Unknown/Declined | 2 ( 0.6%) | 13 ( 0.5%) | 3 ( 1.1%) | 18 ( 0.5%) |  |
| BMI | 29.5 [26.4;33.8] | 30.7 [27.5;35.5] | 31.9 [28.5;36.6] | 30.8 [27.6;35.5] | **<0.01** |
| Maternal obesity (BMI >30) | 95 (47.7%) | 1016 (55.7%) | 127 (64.5%) | 1238 (55.8%) | **<0.01** |
| Obesity classes (BMI) |  |  |  |  | **0.02** |
| - No obesity (BMI<30) | 104 (52.3%) | 807 (44.3%) | 70 (35.5%) | 981 (44.2%) |  |
| - Obesity Class I (BMI 30-34) | 55 (27.6%) | 522 (28.6%) | 57 (28.9%) | 634 (28.6%) |  |
| - Obesity class 2 (BMI35-39) | 23 (11.6%) | 286 (15.7%) | 43 (21.8%) | 352 (15.9%) |  |
| - Obesity class 3 (BMI 40 or over) | 17 ( 8.5%) | 208 (11.4%) | 27 (13.7%) | 252 (11.4%) |  |
| GBS status | 47 (33.1%) | 421 (31.0%) | 49 (33.8%) | 517 (31.5%) | 0.72 |
| SARS-CoV2 status | 28 ( 9.0%) | 149 ( 5.5%) | 10 ( 3.5%) | 187 ( 5.7%) | **0.01** |
| Maternal age (year) | 32.0 [28.0;37.0] | 32.0 [27.0;36.0] | 32.0 [28.5;36.0] | 32.0 [27.0;36.0] |  |
| Delivery mode |  |  |  |  | **0.01** |
| - Cesarean | 90 (28.8%) | 931 (34.4%) | 116 (40.8%) | 1137 (34.4%) |  |
| - Vaginal | 223 (71.2%) | 1774 (65.6%) | 168 (59.2%) | 2165 (65.6%) |  |
| Preeclampsia/PIH | 34 (10.9%) | 394 (14.6%) | 63 (22.2%) | 491 (14.9%) | **<0.01** |
| GDM2 | 40 (12.8%) | 335 (12.4%) | 52 (18.3%) | 427 (12.9%) | **0.02** |
| Category 2 fetal tracing | 72 (23.0%) | 568 (21.0%) | 41 (14.4%) | 681 (20.6%) | **0.02** |
| Placental abruption | 3 ( 1.0%) | 39 ( 1.4%) | 4 ( 1.4%) | 46 ( 1.4%) | 0.79 |
| Oligohydramnios | 7 ( 2.2%) | 64 ( 2.4%) | 3 ( 1.1%) | 74 ( 2.2%) | 0.37 |
| Maternal vascular malperfusion (MVM) |  |  |  |  |  |
| Decidual vasculopathy |  |  |  |  |  |
| - Classic type | 100 (31.9%) | 682 (25.2%) | 68 (23.9%) | 850 (25.7%) | **0.03** |
| - Mixed type | 19 ( 6.1%) | 129 ( 4.8%) | 5 ( 1.8%) | 153 ( 4.6%) | **0.03** |
| - Mural hypertrophy | 20 ( 6.4%) | 211 ( 7.8%) | 33 (11.6%) | 264 ( 8.0%) | **0.04** |
| Infarcts | 23 ( 7.3%) | 166 ( 6.1%) | 21 ( 7.4%) | 210 ( 6.4%) | 0.54 |
| Thrombosis | 70 (22.4%) | 529 (19.6%) | 65 (22.9%) | 664 (20.1%) | 0.24 |
| Fetal vascular malperfusion (FVM) |  |  |  |  |  |
| - Avascular villi | 42 (13.4%) | 322 (11.9%) | 37 (13.0%) | 401 (12.1%) | 0.66 |
| Maternal inflammatory response (MIR) |  |  |  |  |  |
| Acute chorioamnionitis | 120 (38.3%) | 911 (33.7%) | 92 (32.4%) | 1123 (34.0%) | 0.22 |
| Chronic deciduitis (>50/HPF lymph | 100 (31.9%) | 677 (25.0%) | 59 (20.8%) | 836 (25.3%) | **0.01** |
| Chronic villitis | 78 (24.9%) | 546 (20.2%) | 44 (15.5%) | 668 (20.2%) | **0.02** |
| Fetal inflammatory response (FIR) |  |  |  |  |  |
| Acute funisitis / fetal vasculitis | 43 (13.7%) | 347 (12.8%) | 48 (16.9%) | 438 (13.3%) | 0.15 |
| Other placental pathology |  |  |  |  |  |
| Meconium stain | 108 (34.5%) | 857 (31.7%) | 75 (26.4%) | 1040 (31.5%) | 0.09 |
| Subchorionic hematoma (>1.0 cm) | 24 ( 7.7%) | 222 ( 8.2%) | 23 ( 8.1%) | 269 ( 8.1%) | 0.95 |
| MPFD/MFI | 7 ( 2.2%) | 87 ( 3.2%) | 9 ( 3.2%) | 103 ( 3.1%) | 0.64 |
| Lab and other tests |  |  |  |  |  |
| White Blood Cell count (x1000) | 9.9 [ 8.0;12.3] | 9.8 [ 8.2;11.8] | 9.3 [ 7.5;11.1] | 9.7 [ 8.2;11.8] |  |
| Neutrophil differential (%) | 72.6 [68.1;78.3] | 72.5 [67.7;77.4] | 72.5 [67.2;75.9] | 72.5 [67.7;77.4] | 0.43 |
| Lymphocyte differential (%) | 17.9 [13.9;22.6] | 17.8 [14.2;22.1] | 18.5 [14.4;22.5] | 17.9 [14.2;22.2] | 0.47 |
| Body temperature (°C) | 36.7 [36.5;37.0] | 36.7 [36.5;37.0] | 36.8 [36.5;37.0] | 36.7 [36.5;37.0] |  |
| Blood pressure (Systolic) | 125.0 [116.0;132.0] | 125.0 [117.0;135.0] | 130.0 [120.0;141.0] | 126.0 [117.0;135.0] |  |
| Blood pressure (Diastolic) | 77.0 [70.0;81.0] | 77.0 [70.0;84.0] | 80.0 [73.0;88.0] | 77.0 [70.0;84.0] |  |

Abbreviation: IUGR-intrauterine growth restriction; BMI-body mass index; PIH-pregnancy induced hypertension; MPFD/MFI-massive peri-villous fibrinoid deposit/maternal floor infarction.

### 3 Comparison of FPR with fetal birth weight and placental weight

Clinically fetal birth weight has been used to measure neonatal well-being and for clinical assessment of macrosomia or IUGR (FGR). FPR was a poor proxy for assessment of both macrosomia and IUGR (Table 2). In general, FPR significantly correlated with fetal birth weight, but inversely correlated with placental weight (Table 2). Umbilical cord centrality correlated significantly with both fetal birth weight and placental weight but not with FPR. Similarly, preeclampsia/PIH and category 2 fetal heart tracing were significantly associated with FPR and fetal birth weight, but not with placental weight (Table 2). In placental pathology, placental infarcts was significantly associated with fetal birth weight but not with placental weight or FPR. Acute maternal and fetal inflammatory responses and meconium stain of fetal membranes were associated with fetal birth weight and placental weight but not with FPR. Similarly, chronic maternal /fetal inflammatory responses were associated with placental weight or FPR but not with fetal birth weight (Table 2).

**Table 2:** Comparison of FPR, placental and fetal birth weight percentile associated with term pregnancy complications.

| Fetal placental ratio |  |  | Placental weight |  |  | Fetal birth weight |  |  |
| --- | --- | --- | --- | --- | --- | --- | --- | --- |
| Fetal/placental ratio rank | value (% or 95%CI) | p | Placental weight rank | Value (% or 95%CI) | p | Fetal birth weight rank | Value (% or 95%CI) | p |
| - 90% or over | N=313 |  | - 90% or over | N=358 |  | - 90% or over | N=357 |  |
| - 10-89% | N=2705 |  | - 10-89% | N=2741 |  | - 10-89% | N=2804 |  |
| - <10% | N=284 |  | - <10% | N=208 |  | - <10% | N=141 |  |
| Fetal birth birth data characteristics |  |  |  |  |  |  |  |  |
| Fetal birth weight (gram) |  | <0.01 | Fetal birth weight (gram) |  | <0.01 | Fetal birth weight (gram) |  | <0.01 |
| - 90% or over | 3280 [2940;3655] |  | - 90% or over | 3870 [3560;4160] |  | - 90% or over | 4130 [4010;4330] |  |
| - 10-89% | 3310 [2990;3630] |  | - 10-89% | 3280 [2990;3570] |  | - 10-89% | 3260 [2980;3520] |  |
| - <10% | 3150 [2880;3470] |  | - <10% | 2635 [2395;2880] |  | - <10% | 2290 [2150;2380] |  |
| Macrosomia (>4000 g) |  | 0.08 | Macrosomia (>4000 g) |  | <0.01 | Macrosomia (>4000 g) |  | <0.01 |
| - 90% or over | 29 ( 9.3%) |  | - 90% or over | 140 (39.1%) |  | - 90% or over | 277 (77.6%) |  |
| - 10-89% | 234 ( 8.7%) |  | - 10-89% | 137 ( 5.0%) |  | - 10-89% | 0 ( 0.0%) |  |
| - <10% | 14 ( 4.9%) |  | - <10% | 0 ( 0.0%) |  | - <10% | 0 ( 0.0%) |  |
| IUGR (<10%) |  | 0.31 | IUGR (weight less than 10%) |  | <0.01 | IUGR (<10%) |  | <0.01 |
| - 90% or over | 19 ( 6.1%) |  | - 90% or over | 0 ( 0.0%) |  | - 90% or over | 0 ( 0.0%) |  |
| - 10-89% | 122 ( 4.5%) |  | - 10-89% | 94 ( 3.4%) |  | - 10-89% | 100 ( 3.6%) |  |
| - <10% | 10 ( 3.5%) |  | - <10% | 57 (27.4%) |  | - <10% | 51 (36.2%) |  |
| Placental weight (gram) |  | <0.01 | Placental weight (trimmed, gram) |  | <0.01 | Placental weight (gram) |  | <0.01 |
| - 90% or over | 354 [314;396] |  | - 90% or over | 655 [628;692] |  | - 90% or over | 595 [535;664] |  |
| - 10-89% | 467 [413;530] |  | - 10-89% | 459 [410;517] |  | - 10-89% | 457 [403;522] |  |
| - <10% | 605 [548;665] |  | - <10% | 307 [284;322] |  | - <10% | 341 [284;382] |  |
| Cord insertion to edge (cm) |  | 0.31 | Insertion site to edge (cm)(centrality) |  | <0.01 | Insertion site to edge (cm)(centrality) |  | <0.01 |
| - 90% or over | 4.0 [ 3.0; 5.3] |  | - 90% or over | 4.3 [ 3.0; 5.8] |  | - 90% or over | 5.0 [ 3.5; 6.0] |  |
| - 10-89% | 4.3 [ 3.0; 5.4] |  | - 10-89% | 4.3 [ 3.0; 5.4] |  | - 10-89% | 4.2 [ 3.0; 5.3] |  |
| - <10% | 4.2 [ 3.0; 5.3] |  | - <10% | 3.8 [ 2.5; 4.8] |  | - <10% | 3.8 [ 2.5; 4.5] |  |
| Maternal data characteristics |  |  |  |  |  |  |  |  |
| Maternal obesity (BMI >30) |  | <0.01 | Maternal obesity (BMI >30) |  | <0.01 | Maternal obesity (BMI >30) |  | <0.01 |
| - 90% or over | 95 (47.7%) |  | - 90% or over | 169 (68.4%) |  | - 90% or over | 176 (73.6%) |  |
| - 10-89% | 1016 (55.7%) |  | - 10-89% | 1012 (55.1%) |  | - 10-89% | 1011 (53.6%) |  |
| - <10% | 127 (64.5%) |  | - <10% | 60 (43.2%) |  | - <10% | 51 (54.3%) |  |
| SARS-CoV2 status |  | 0.01 | SARS-CoV2 status |  | 0.08 | SARS-CoV2 status |  | 0.45 |
| - 90% or over | 28 ( 9.0%) |  | - 90% or over | 11 ( 3.1%) |  | - 90% or over | 15 ( 4.2%) |  |
| - 10-89% | 149 ( 5.5%) |  | - 10-89% | 163 ( 5.9%) |  | - 10-89% | 164 ( 5.9%) |  |
| - <10% | 10 ( 3.5%) |  | - <10% | 13 ( 6.2%) |  | - <10% | 8 ( 5.7%) |  |
| Delivery mode |  | **0.01** | Delivery mode |  | **<0.01** | Delivery mode |  | **0.02** |
| - Cesarean |  |  | - Cesarean |  |  | - Cesarean |  |  |
| - 90% or over | 90 (28.8%) |  | - 90% or over | 171 (47.8%) |  | - 90% or over | 146 (40.9%) |  |
| - 10-89% | 931 (34.4%) |  | - 10-89% | 901 (32.9%) |  | - 10-89% | 938 (33.5%) |  |
| - <10% | 116 (40.8%) |  | - <10% | 67 (32.2%) |  | - <10% | 53 (37.6%) |  |
| Preeclampsia/PIH |  | **<0.01** | Preeclampsia/PIH |  | 0.19 | Preeclampsia/PIH |  | **<0.01** |
| - 90% or over | 34 (10.9%) |  | - 90% or over | 53 (14.8%) |  | - 90% or over | 46 (12.9%) |  |
| - 10-89% | 394 (14.6%) |  | - 10-89% | 400 (14.6%) |  | - 10-89% | 409 (14.6%) |  |
| - <10% | 63 (22.2%) |  | - <10% | 40 (19.2%) |  | - <10% | 36 (25.5%) |  |
| GDM2 |  | **0.02** | GDM2 |  | **0.01** | GDM2 |  | **0.01** |
| - 90% or over | 40 (12.8%) |  | - 90% or over | 64 (17.9%) |  | - 90% or over | 64 (17.9%) |  |
| - 10-89% | 335 (12.4%) |  | - 10-89% | 337 (12.3%) |  | - 10-89% | 348 (12.4%) |  |
| - <10% | 52 (18.3%) |  | - <10% | 26 (12.5%) |  | - <10% | 15 (10.6%) |  |
| Category 2 fetal tracing |  | **0.02** | Category 2 fetal tracing |  | 0.09 | Category 2 fetal tracing |  | **0.01** |
| - 90% or over | 72 (23.0%) |  | - 90% or over | 58 (16.2%) |  | - 90% or over | 52 (14.6%) |  |
| - 10-89% | 568 (21.0%) |  | - 10-89% | 582 (21.2%) |  | - 10-89% | 605 (21.6%) |  |
| - <10% | 41 (14.4%) |  | - <10% | 42 (20.2%) |  | - <10% | 24 (17.0%) |  |
| **Placental pathology** |  |  |  |  |  |  |  |  |
| **Maternal vascular malperfusion (MVM)** |  |  |  |  |  |  |  |  |
| Infarcts |  | 0.54 | Infarcts |  | 0.24 | Infarcts |  | **<0.01** |
| - 90% or over | 23 ( 7.3%) |  | - 90% or over | 21 ( 5.9%) |  | - 90% or over | 17 ( 4.8%) |  |
| - 10-89% | 166 ( 6.1%) |  | - 10-89% | 172 ( 6.3%) |  | - 10-89% | 175 ( 6.2%) |  |
| - <10% | 21 ( 7.4%) |  | - <10% | 19 ( 9.1%) |  | - <10% | 18 (12.8%) |  |
| **Inflammatory/Infectious** |  |  |  |  |  |  |  |  |
| **Maternal inflammatory response (MIR)** |  |  |  |  |  |  |  |  |
| Acute chorioamnionitis |  | 0.22 | Acute chorioamnionitis |  | **<0.01** | Acute chorioamnionitis |  | **<0.01** |
| - 90% or over | 120 (38.3%) |  | - 90% or over | 123 (34.4%) |  | - 90% or over | 130 (36.4%) |  |
| - 10-89% | 911 (33.7%) |  | - 10-89% | 955 (34.8%) |  | - 10-89% | 967 (34.5%) |  |
| - <10% | 92 (32.4%) |  | - <10% | 46 (22.1%) |  | - <10% | 26 (18.4%) |  |
| Chronic deciduitis (>50/HPF ) |  | **0.01** | Chronic deciduitis (>50/HPF) |  | **0.01** | Chronic deciduitis (>50/HPF ) |  | 0.20 |
| - 90% or over | 100 (31.9%) |  | - 90% or over | 72 (20.1%) |  | - 90% or over | 77 (21.6%) |  |
| - 10-89% | 677 (25.0%) |  | - 10-89% | 700 (25.5%) |  | - 10-89% | 720 (25.7%) |  |
| - <10% | 59 (20.8%) |  | - <10% | 64 (30.8%) |  | - <10% | 39 (27.7%) |  |
| Chronic villitis |  | **0.02** | Chronic villitis |  | 0.10 | Chronic villitis |  | 0.30 |
| - 90% or over | 78 (24.9%) |  | - 90% or over | 59 (16.5%) |  | - 90% or over | 63 (17.6%) |  |
| - 10-89% | 546 (20.2%) |  | - 10-89% | 561 (20.5%) |  | - 10-89% | 572 (20.4%) |  |
| - <10% | 44 (15.5%) |  | - <10% | 49 (23.6%) |  | - <10% | 33 (23.4%) |  |
| **Fetal inflammatory response (FIR)** |  |  |  |  |  |  |  |  |
| Acute funisitis / fetal vasculitis |  | 0.15 | Acute funisitis /fetal vasculitis |  | **0.01** | Acute funisitis / fetal vasculitis |  | **0.03** |
| - 90% or over | 43 (13.7%) |  | - 90% or over | 55 (15.4%) |  | - 90% or over | 54 (15.1%) |  |
| - 10-89% | 347 (12.8%) |  | - 10-89% | 370 (13.5%) |  | - 10-89% | 375 (13.4%) |  |
| - <10% | 48 (16.9%) |  | - <10% | 14 ( 6.7%) |  | - <10% | 9 ( 6.4%) |  |
| **Other placental pathology** |  |  |  |  |  |  |  |  |
| Meconium stain |  | 0.09 | Meconium stain |  | **0.04** | Meconium stain |  | **<0.01** |
| - 90% or over | 108 (34.5%) |  | - 90% or over | 113 (31.6%) |  | - 90% or over | 121 (33.9%) |  |
| - 10-89% | 857 (31.7%) |  | - 10-89% | 878 (32.0%) |  | - 10-89% | 897 (32.0%) |  |
| - <10% | 75 (26.4%) |  | - <10% | 49 (23.6%) |  | - <10% | 22 (15.6%) |  |
| MPFD/MFI |  | 0.64 | MPFD/MFI |  | 0.49 | MPFD/MFI |  | **0.02** |
| - 90% or over | 7 ( 2.2%) |  | - 90% or over | 9 ( 2.5%) |  | - 90% or over | 10 ( 2.8%) |  |
| - 10-89% | 87 ( 3.2%) |  | - 10-89% | 85 ( 3.1%) |  | - 10-89% | 83 ( 3.0%) |  |
| - <10% | 9 ( 3.2%) |  | - <10% | 9 ( 4.3%) |  | - <10% | 10 ( 7.1%) |  |

Abbreviation: IUGR-intrauterine growth restriction; BMI-body mass index; PIH - pregnancy induced hypertension; MPFD/MFI-massive peri-villous fibrinoid deposit/maternal floor infarction.

In summary (Table 3), the ethnic White population, SARS-CoV2 status, category 2 fetal tracing, classic and mixed type decidual vasculopathy, and chronic maternal inflammatory responses were significantly associated with high placental efficiency (measured as FPR 90% or over). The ethnic Asian, Black, Hispanic races, increased Cesarean section rate, preeclampsia, GDM2 and mural artery hypertrophy were associated with low placental efficiency (FPR <10%).

**Table 3:** Placental efficiency and clinical complications.

| **HPE (FPR 90% or over)**<br>(N=313) | **LPE (FPR <10%)**<br>(N=284) | **p value** |
| --- | --- | --- |
|  | **Race/ethnicity** | **<0.01** |
|  | Asian |  |
|  | Black |  |
|  | Hispanic |  |
| White |  |  |
| SARS-CoV2 status |  | **0.01** |
|  | **Clinical complications** |  |
|  | Cesarean | 0.01 |
|  | Preeclampsia/PIH | **<0.01** |
|  | GDM2 | **0.02** |
| Category 2 fetal tracing |  | **0.02** |
|  | **Maternal vascular malperfusion** |  |
|  | **Decidual vasculopathy** |  |
| Classic type |  | **0.03** |
| Mixed type |  | **0.03** |
|  | Mural hypertrophy | **0.04** |
|  | **Maternal inflammatory responses** |  |
| Chronic deciduitis (>50/HPF lymphocytes) |  | **0.01** |
| Chronic villitis |  | **0.02** |

## Discussion

Our data indicated that placental efficiency is associated with different clinical complications of pregnancy as well as placental pathology. Maternal characteristics, pregnancy complications and placental pathology can be stratified based on the placental efficiency as measured by the FPR. Ethnic Asian, Black, and Hispanic mothers, increased Cesarean section rate, preeclampsia and GDM2 were statistically significantly associated with low placental efficiency, whereas ethnic White, SARS-CoV2 status, maternal decidual vasculopathy (both classic type and mixed type) as well as maternal inflammatory responses were associated with high placental efficiency. These data demonstrated that the clinically important maternal conditions or environmental factors affect the fetal weight and placental weight unequally, and some conditions affect the growth of fetus more, and others on placenta more [18]. Surprisingly, SARS-CoV2 status was statistically associated only with high placental efficiency, similar to other maternal inflammatory responses such as chronic deciduitis or chronic villitis, but not with fetal birth weight or placental weight alone [19]. It is noteworthy that GDM2 in our data was associated with low placental efficiency, rather than high placental efficiency, and GDM2 is known to be associated with macrosomia or large for gestational age fetuses (LGA) and large placentas [2, 20]. Our data suggests that GDM2 affects placental growth more than fetal growth, although both fetal and placental growth were increased [21, 22]. Animal studies with IGFs and receptors showed that the growth pattern of fetal tissue and/or placental tissues in mid-gestation was different from that at term or near term [23-25]. Increased IGFs in maternal circulation in mid-gestation increased both the fetal weight and placental weight, but increased IGFs at term pregnancy only increased the fetal weight but not placental weight in guinea pigs [23-25]. Preeclampsia/PIH was known to be associated with low placental efficiency and fetal growth restriction [18]. However, term preeclampsia/PIH in our data was associated with low placental efficiency and low fetal weight but not with low placental weight, and the term preeclampsia/PIH was different from preterm or early onset preeclampsia as previously described [26].

Many other maternal factors including maternal racial/ethnic minorities, nutritional health, social economic status, emotional health and stress as well as marital status can influence fetal birth weight and placental weight unequally [27-30]. Marital status affected fetal birth weight but not placental weight, leading to differences in placental efficiency. Social emotional health / maternal stress or marital status affected the maternal/fetal physical health and placental pathology mainly through glucocorticoid hormones, and there were significant associations between prenatal glucocorticoids and neonatal complications and adult life [31, 32]. The effects of glucocorticoids on fetal and placental growth again depended on the gestational age in animal studies [33-35].

## Conclusions

Placental efficiency measured as FPR serves as a different parameter from fetal birth weight and placental weight alone in assessment of fetal wellbeing immediately after birth or later in life. Pregnancy complications can be stratified based on the placental efficiency and FPR can be incorporated into clinical information system to help assess the neonatal health and disease status.

## Data Availability

All data produced in the present work are contained in the manuscript.

## References

1. Cunningham, F.G., Williams obstetrics. 25th edition. ed. 2018, New York: McGraw-Hill. xvi, 1328 pages.

2. Benirschke, K., G.J. Burton, and R.N. Baergen Pathology of the Human Placenta. 6th ed. 2012: Springer.

3. Little, W.A., The significance of placental/fetal weight ratios. Am J Obstet Gynecol, 1960. 79: p. 134–7.

4. Risnes, K.R., et al., Placental weight relative to birth weight and long-term cardiovascular mortality: findings from a cohort of 31,307 men and women. Am J Epidemiol, 2009. 170(5): p. 622–31.

5. Yampolsky, M., et al., Centrality of the umbilical cord insertion in a human placenta influences the placental efficiency. Placenta, 2009. 30(12): p. 1058–64.

6. Hayward, C.E., et al., Placental Adaptation: What Can We Learn from Birthweight:Placental Weight Ratio? Front Physiol, 2016. 7: p. 28.

7. Fowden, A.L., et al., Placental efficiency and adaptation: endocrine regulation. J Physiol, 2009. 587(Pt 14): p. 3459–72.

8. Barker, D.J., The fetal and infant origins of adult disease. BMJ, 1990. 301(6761): p. 1111.

9. Thornburg, K.L. and N. Marshall, The placenta is the center of the chronic disease universe. Am J Obstet Gynecol, 2015. 213(4 Suppl): p. S14–20.

10. Barker, D.J., et al., The placental origins of sudden cardiac death. Int J Epidemiol, 2012. 41(5): p. 1394–9.

11. Barker, D.J. and K.L. Thornburg, Placental programming of chronic diseases, cancer and lifespan: a review. Placenta, 2013. 34(10): p. 841–5.

12. Burton, G.J., A.L. Fowden, and K.L. Thornburg, Placental Origins of Chronic Disease. Physiol Rev, 2016. 96(4): p. 1509–65.

13. Coan, P.M., et al., Adaptations in placental nutrient transfer capacity to meet fetal growth demands depend on placental size in mice. J Physiol, 2008. 586(18): p. 4567–76.

14. Echternkamp, S.E., Relationship between placental development and calf birth weight in beef cattle. Animal Reproduction Science, 1993. 32: p. 1–13.

15. Khong, T.Y., et al., Sampling and Definitions of Placental Lesions: Amsterdam Placental Workshop Group Consensus Statement. Arch Pathol Lab Med, 2016. 140(7): p. 698–713.

16. Langston, C., et al., Practice guideline for examination of the placenta: developed by the Placental Pathology Practice Guideline Development Task Force of the College of American Pathologists. Arch Pathol Lab Med, 1997. 121(5): p. 449–76.

17. Baergen, R.N., Indications for submission and macroscopic examination of the placenta. APMIS, 2018. 126(7): p. 544–550.

18. Burton, G.J. and E. Jauniaux, Pathophysiology of placental-derived fetal growth restriction. Am J Obstet Gynecol, 2018. 218(2S): p. S745–S761.

19. Schwartz, D.A. and D. Morotti, Placental Pathology of COVID-19 with and without Fetal and Neonatal Infection: Trophoblast Necrosis and Chronic Histiocytic Intervillositis as Risk Factors for Transplacental Transmission of SARS-CoV-2. Viruses, 2020. 2(11).

20. Zhang, P., et al., Placental pathology associated with maternal age and maternal obesity in singleton pregnancy. J Matern Fetal Neonatal Med, 2022: p. 1–10.

21. Tanaka, K., et al., Increased maternal insulin resistance promotes placental growth and decreases placental efficiency in pregnancies with obesity and gestational diabetes mellitus. J Obstet Gynaecol Res, 2018. 44(1): p. 74–80.

22. Weissgerber, T.L. and L.M. Mudd, Preeclampsia and diabetes. Curr Diab Rep, 2015. 15(3): p. 9.

23. Aykroyd, B.R.L., S.J. Tunster, and A.N. Sferruzzi-Perri, Igf2 deletion alters mouse placenta endocrine capacity in a sexually dimorphic manner. J Endocrinol, 2020. 246(1): p. 93–108.

24. Harris, L.K., et al., IGF2 actions on trophoblast in human placenta are regulated by the insulin-like growth factor 2 receptor, which can function as both a signaling and clearance receptor. Biol Reprod, 2011. 84(3): p. 440–6.

25. Coan, P.M., et al., Disproportional effects of Igf2 knockout on placental morphology and diffusional exchange characteristics in the mouse. J Physiol, 2008. 586(20): p. 5023–32.

26. Zhang, P. and N. Shama, Early and late onset preeclampsia associated with different placental pathology and clinical risk characteristics. MedRxiv, 2022.

27. Zhang, P., et al., Potential association between marital status and maternal and neonatal complications and placental pathology in singleton pregnancy. Reproductive Medicine, 2023. 4: p. 28–33.

28. Zhang, P., et al., Differences in Prevalence of Pregnancy Complications and Placental Pathology by Race and Ethnicity in a New York Community Hospital. JAMA Netw Open, 2022. 5(5): p. e2210719.

29. Dahlerup, B.R., et al., Maternal stress and placental function, a study using questionnaires and biomarkers at birth. PLoS One, 2018. 13(11): p. e0207184.

30. Send, T.S., et al., Telomere Length in Newborns is Related to Maternal Stress During Pregnancy. Neuropsychopharmacology, 2017. 42(12): p. 2407–2413.

31. Laugesen, K., et al., Prenatal exposure to glucocorticoids and the prevalence of overweight or obesity in childhood. Eur J Endocrinol, 2022. 186(4): p. 429–440.

32. Laugesen, K., et al., In utero exposure to glucocorticoids and risk of anxiety and depression in childhood or adolescence. Psychoneuroendocrinology, 2022. 141: p. 105766.

33. Ain, R., et al., Phenotypic analysis of the rat placenta. Methods Mol Med, 2006. 121: p. 295–313.

34. Ain, R., L.N. Canham, and M.J. Soares, Dexamethasone-induced intrauterine growth restriction impacts the placental prolactin family, insulin-like growth factor-II and the Akt signaling pathway. J Endocrinol, 2005. 185(2): p. 253–63.

35. Ain, R., et al., A prolactin family paralog regulates reproductive adaptations to a physiological stressor. Proc Natl Acad Sci U S A, 2004. 101(47): p. 16543–8.

